# Estimates of Single Dose and Full Dose BNT162b2 Vaccine Effectiveness among USAF Academy cadets, 1 Mar - 1 May 2021

**DOI:** 10.1101/2021.07.28.21261138

**Authors:** Douglas P. Wickert, Erin A. Almand, Kevin J. Baldovich, Christopher A. Cullenbine, Odaro J. Huckstep, Joseph W. Rohrer, John C. Sitko, J. Jordan Steel, Steven C.M. Hasstedt

## Abstract

Beginning in early March 2021 and continuing through May 2021, the USAF Academy began vaccinating cadets for protection against the SARS-CoV-2 virus with the BNT162b2 (Pfizer-BioNTech) mRNA vaccine. During this period, vaccination of the almost 4200 cadet population increased from 3% to 85% and prevalence of COVID-19 in the cadet population was constant at approximately 0.4% as indicated by weekly surveillance testing. In this study, vaccine effectiveness at preventing infection is estimated by comparing infection risk as a function of time since vaccination. A statistically significant four-fold reduction in infection risk was observed 14 days after the first vaccine dose and an eleven-fold reduction in infection risk was observed in fully vaccinated cadets. Overall, the Pfizer-BioNTech vaccine was 91% (95% confidence interval = 55-99%) effective at preventing infection in healthy young adults (17-26 years of age) in a university setting and military training environment.

## Methods

The USAF Academy is a university located on a military base in Colorado Springs, CO, with a student body of approximately 4200 cadets between the ages of 17 and 26 years (the vast majority of cadets are between 18 and 22 years of age). All cadets live in two large dormitories on campus and attended in-person academic, military, and athletic training during the study period. Vaccination of cadets was voluntary as the Pfizer-BioNTech vaccine was approved under Emergency Use Authorization (EUA) and was conducted in accordance with DoD vaccination priority. Weekly surveillance testing using the Sofia SARS Antigen Fluorescent Immunoassay (FIA) (Quidel Corporation) provided infection point prevalence estimates throughout the spring semester, including the entire study period. Asymptomatic individuals identified during surveillance testing and symptomatic individuals reporting to the cadet clinic for testing received RT-PCR based tests using the GeneXpert Xpress SARS-CoV-2 assay (Cepheid Innovation) approved for diagnostic use under EUA.

**Table:** Vaccination cohort person-days, number of SARS-CoV-2 infections, infection rate, and vaccine effectiveness among USAF Academy cadets, 1 Mar – 1 May 2021

| Cohort<sup>(1)</sup> | Person-days | # SARS-CoV-2 infections | Infection rate (per 10k person-days) (95% CI)<sup>(2)</sup> | Vaccine Effectiveness<sup>(3)</sup> (95% CI) |
| --- | --- | --- | --- | --- |
| unvaccinated | 127,893 | 112 | 8.76 (7.2-10.6) | -- |
| week 1 (0-6 days) | 23,856 | 17 | 7.12 (4.3-11.7) | 19% (0-59%) |
| week 2 (7-13 days) | 23,454 | 11 | 4.69 (2.5-8.7) | 46% (0-77%) |
| week 3 (14-20days) | 22,834 | 5 | 2.19 (0.8-5.4) | 75% (25-92%) |
| week 4 (21-27 days)<sup>(4)</sup> | 19,621 | 4 | 2.04 (0.7-5.6) | 77% (22-94%) |
| week 5 (28-34 days) | 6,919 | 0 | 0 (0-6.9) | -- |
| fully vaccinated<sup>(5)</sup> | 24,791 | 2 | 0.81 (0.1-3.3) | 91% (55-99%) |
| total<sup>(6)</sup> | 255,468 | 151 | -- | -- |

<sup>1</sup>: acquired immunity from previous infections is not accounted for cohort description
<sup>2</sup>: 95% confidence interval is the Wilson interval
<sup>3</sup>: vaccine effectiveness = (unvaccinated attack rate – vaccinated attack rate) / (unvaccinated attack rate)
<sup>4</sup>: in most cases, 2<sup>nd</sup> dose of BNT162b2 (Pfizer-BioNTech) vaccine given on day 21
<sup>5</sup>: fully vaccinated defined as 14 days after the second dose
<sup>6</sup>: total = (4188 cadets)(61 days) = 255,468 person-days

A consistent infection point prevalence among the cadet population during the 61 days between 1 March and 1 May permits a natural experiment of post-vaccination immunity development and an estimate of vaccine effectiveness in a healthy, 17-26 year-old demographic. Infection risk as a function of vaccination time status was determined by comparing the total person-days within the observation period and the total infection count, per 10,000 person-days for each vaccination group (*Table*). These rates were calculated separately for those individuals with partial vaccination. Statistical significance was determined using the Fisher’s Exact test with each partially and fully vaccinated group compared to unvaccinated infection rates. The odds ratio was calculated to determine the likelihood that a vaccinated cadet becomes infected compared to an unvaccinated cadet. All statistics were performed using the SciPy-Stats Python library.

## Results

The vaccine effectiveness analytic study began on 1 March when less than 3% of cadets had received any dose of vaccine and concluded on 1 May when infection point prevalence dropped below 0.4%. By 1 May, 85% of cadets had received at least a first dose of the BNT162b2 vaccine, 71% of cadets had received a second dose of the BNT162b2 vaccine, and 36% of cadets were fully vaccinated (defined as 14 days after the second dose per for the CDC definition at the time) (*Figure 1, top*). During this period, point prevalence of COVID-19 infections averaged 0.42% of the cadet population (95% confidence interval range of 0.2-0.8%) based on weekly surveillance testing of the cadet wing (*Figure 1, bottom*). Weekly surveillance test volume ranged from 100% to 50% of the cadet population (*Figure 1, bottom*). This study was restricted to the 61 days between 1 March and 1 May because infection prevalence was greater than 0.8% before 1 March and decreased to 0.1% after 1 May representing a significant change in underlying exposure risk.

**Figure 1:**
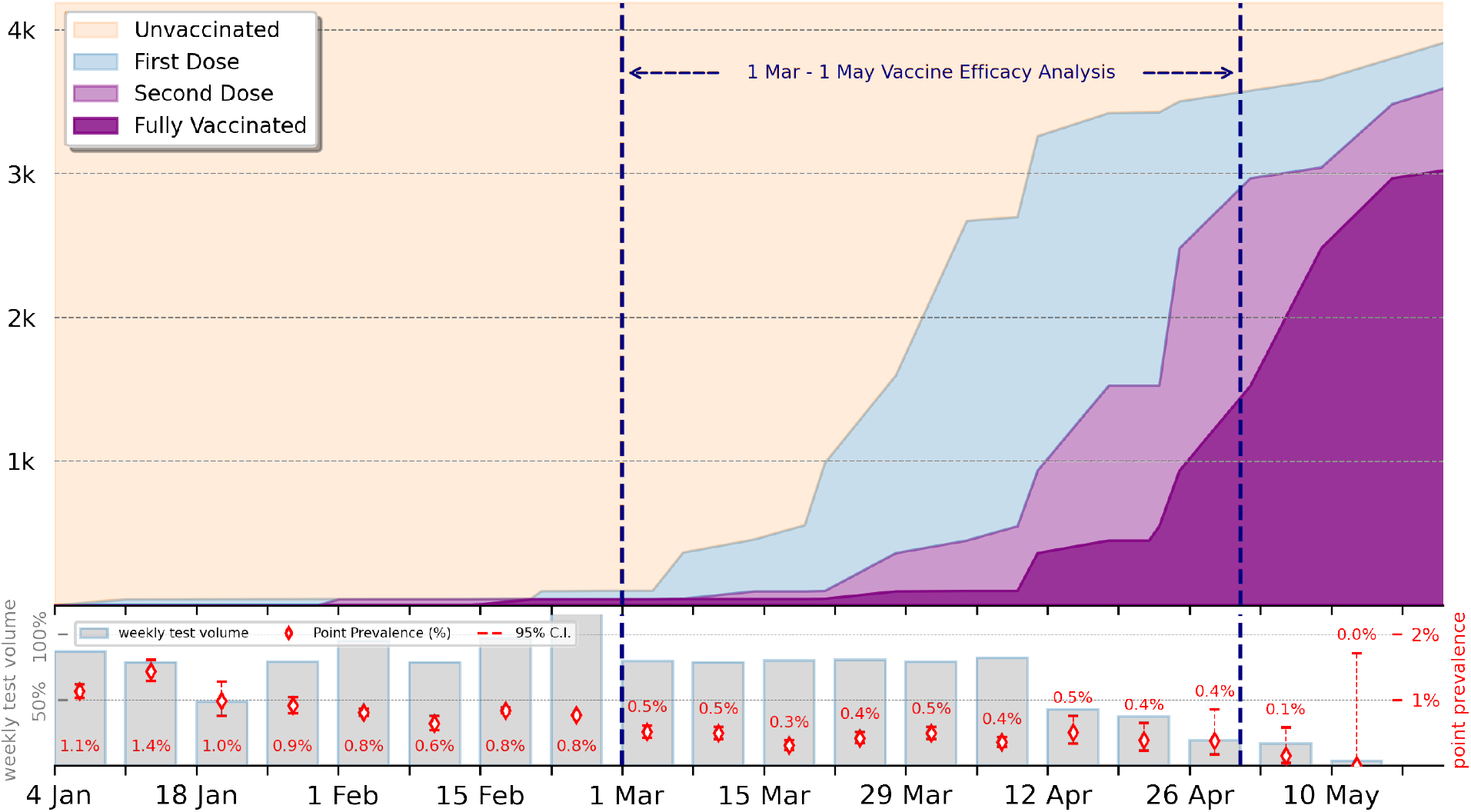
(top) USAF Academy cadet vaccination rollout; (bottom) weekly surveillance test volume and point prevalence estimate (weekly percentage value given) with 95% confidence interval (based on random, hypergeometric sample of the cadet population)

A total of 151 COVID-19 infections were recorded between 1 March and 1 May: 112 cases in unvaccinated cadets, 37 cases in partially vaccinated cadets, and 2 cases in fully vaccinated cadets (*Table*). *Figure 2* provides detail by day of post-vaccination for each of the 37 partially vaccinated cases and the 2 fully vaccinated cases. Summing the vaccination status for all cadets in week 1 (0-6 days), week 2 (7-13 days), week 3 (14-20 days), week 4 (21-27 days)^*^, and week 5 (28-34 days) after vaccination provides the total number of per-person vaccination days in each of the groups. *Figure 3* compares the infection rate per 10k person-days for each of the vaccination week cohorts with the unvaccinated cohort during the period of 1 March to 1 May when infection prevalence was approximately constant across the population.

**Figure 2:**
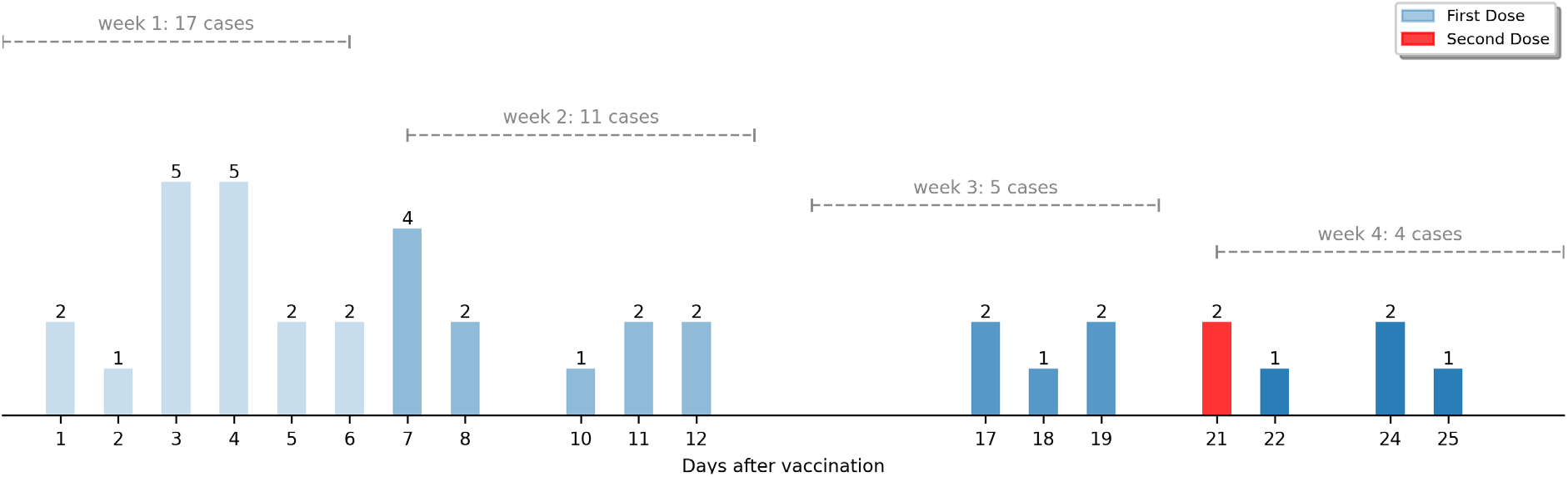
USAF Academy cadet post-vaccination infections (1 Mar – 1 May 2021); the two vaccine-breakthrough cases both occurred 21 days after the second dose

**Figure 3:**
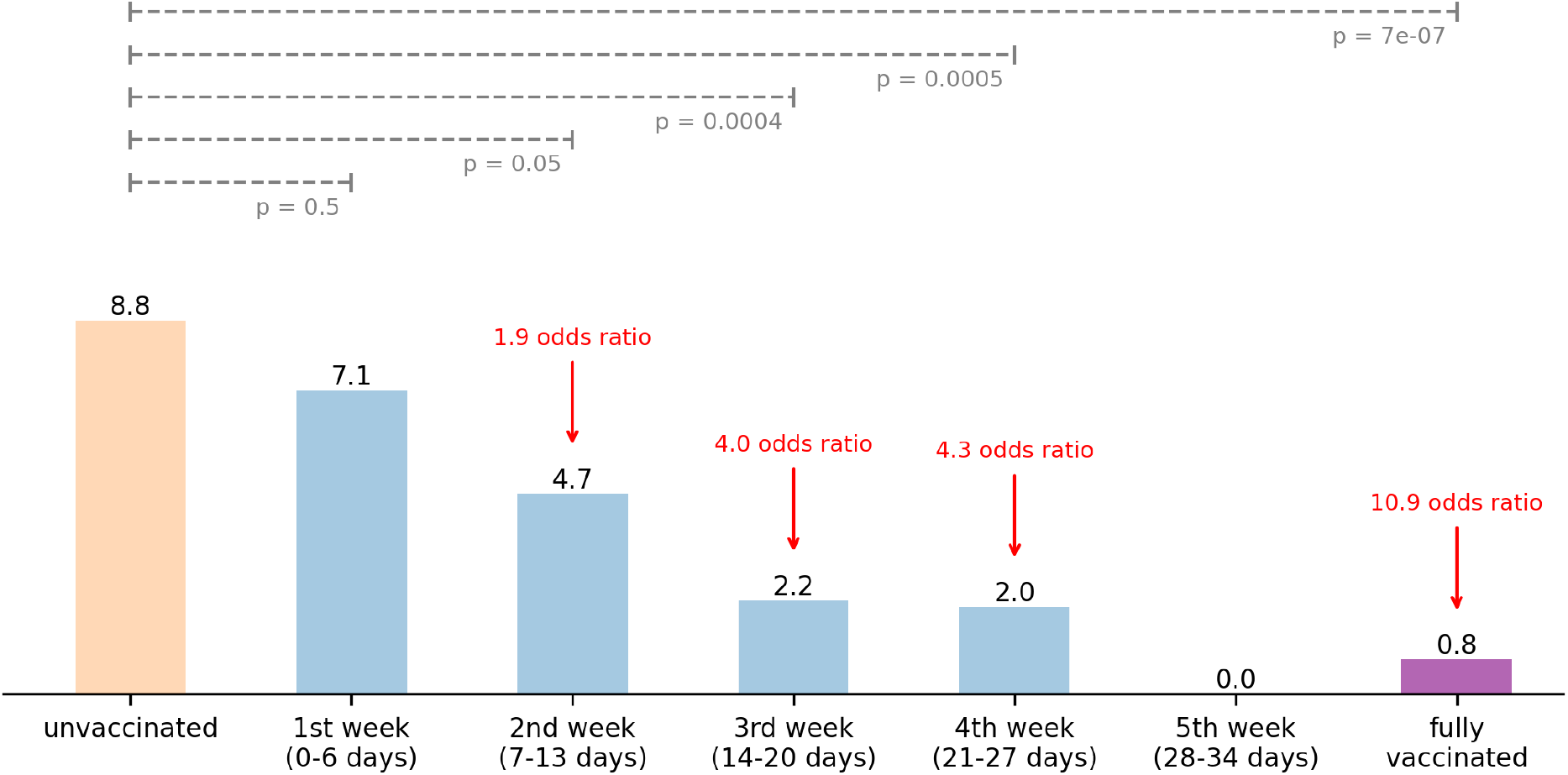
USAF Academy cadet post-vaccination infection rate (per 10k person-days); *p-*values based on Fisher’s exact test compared to unvaccinated group

A statistically modest reduction of 1.9 in infection risk appeared by the second week after administration of a first dose of the BNT162b2 mRNA vaccine (*p* = 0.05)^†^. By the start of the third week, *i*.*e*., 14 days after the first dose, a statistically significant four-fold reduction of infection risk was observed (*p* = 0.0004). Immunity continued to increase with time. A statistically significant eleven-fold reduction of infection risk was observed in fully vaccinated cadets (*p* = 7E-7). These data are consistent with observations of vaccination of healthcare workers in previous vaccine effectiveness studies in the UK [1] and the US [2].

## Discussion

This single-site analysis of vaccination effectiveness found the Pfizer-BioNTech BNT162b2 mRNA vaccine to be highly effective (91%) (95% confidence interval = 55-99%) against symptomatic and asymptomatic COVID-19 amongst a healthy population of 17-26 year olds with no known comorbidities. Infections were tracked based on days post vaccination, with vaccine distribution scheduled to ensure the second-dose of the vaccine occurred at 21 days post initial dose in almost all cadets. Positive infections during the study period were identified based on surveillance and symptomatic testing. After 14-days post initial vaccine dose, a four-fold reduction in infection risk was observed (*Figure 3*). Among fully xzvaccinated cadets, an eleven-fold reduction in infection risk was observed (*Figure 3*). With every week post-vaccination, there is a statically significant drop in infection risk compared to unvaccinated individuals.

Limitations to the present study include potential variations in SARS-CoV-2 exposure risk during the time period. Weekly surveillance testing indicated approximately constant prevalence of COVID-19 during the analysis period, but surveillance testing was done with Sofia SARS Antigen Fluorescent Immunoassay which has a known low sensitivity (41%) when used for screening of asymptomatic persons [3]. A variation in exposure risk during the analysis period would alter infection risk as more of the cadet population was vaccinated over time. A second limitation is that the present analysis does not account for potential acquired immunity due to prior COVID-19 infections. Approximately 10-15% of the cadet population had previously tested positive for SARS-CoV-2 infection since May 2020, but since estimates for durable immunity are not available and cadets were vaccinated regardless of prior infection status, we did not account for prior infection in determining infection risk. A third limitation are prominent SARS-CoV-2 strains present during the study period. Based on random sequencing (approximately one-quarter of positive infection samples were sequenced), the B.1.429 strain was predominant (76%) at the beginning of the study period. The B.1.1.7 strain increased in prevalence during the study period. Both of two the breakthrough infections were B.1.1.7 variants. The Pfizer-BioNTech vaccine is effective against all known strains during the study period.

The continuously developing immunity as a function of time after vaccination and the findings of overall vaccine effectiveness in this study demonstrate vaccine efficacy in university and military settings. These results reinforce the CDC’s recommendation of vaccination against COVID-19 for all eligible persons.

### Summary

#### What is already known about this topic?

The BNT162b2 (Pfizer-BioNTech) mRNA vaccine has been shown to be effective in preventing symptomatic SARS-CoV-2 infection in randomized placebo-controlled Phase III trials. Interim estimates have shown 80% vaccine effectiveness ≥14 days after first dose and 90% effectiveness for fully vaccinated against symptomatic and asymptomatic infection.

#### What is added by this report?

Under real-world conditions in a university setting and in a military training environment, the BNT162b2 (Pfizer-BioNTech) mRNA vaccine is better than 90% effective at preventing symptomatic and asymptomatic infections in young, healthy adults. Reduction in infection risk as early as the second week after a first dose was observed in partially immunized individuals. This increases to a four-fold reduction in infection risk ≥14 days after a first dose.

#### What are the implications for public health practice?

The Pfizer-BioNTech mRNA vaccine is effective at preventing both symptomatic and asymptomatic SARS-CoV-2 infection in university and military training environments. COVID-19 vaccination is recommended for all eligible persons.

## Data Availability

All data is available. Data has been anonymized to remove PII and HIPAA from the data set.

## Acknowledgments

The leadership and personnel of the 10th Medical Group, United States Air Force Academy, Colorado.

## Footnotes

* In most, but not all cases, cadets received a second dose of BNT162b2 vaccine on day 21

† All *p*-values based on a Fisher’s exact test between the given week cohort and unvaccinated cohort; the SciPy-Stats Python library was used for all statistical calculations

## Notes

### Competing Interest Statement

The authors have declared no competing interest.

### Author Declarations

IRB was not required. 10th Medical Group, USAF Academy, is the designated medical ethics oversight body at the USAF Academy. The 10 MDG HIPAA compliance officer referred the vaccination effectiveness study plan in Feb 2021 and granted permission to proceed with analysis. Data was collected as part of the declared public health emergency and anonymized for the purposes of this study.

